# Optimal lockdown strategies for SARS-CoV2 mitigation— an Indian perspective

**DOI:** 10.1101/2020.07.31.20165662

**Authors:** Anagh Pathak, Varun Madan Mohan, Arpan Banerjee

**Affiliations:** National Brain Research Centre, Manesar, Gurgaon 122052, India

**Keywords:** Lockdown, Optimal temporal window, Infection, SARS-CoV2, Viral spread, Intervention, Phase dependence

## Abstract

We sought to identify optimal temporal windows for lockdown-based mitigation strategies on infectious disease spreads. An age-structured multi-compartmental Susceptible-Infected-Recovered (SIR) model was used to estimate infection spreads under parametric variation of lockdown intensity and duration from the data of SARS-CoV2 cases in India between January to July, 2020. The resulting parameter values were used to simulate lockdown outcomes for a wide range of start times and durations. Lockdowns were simulated as intervention strategies that modified weights assigned to social contact matrices for work, school and other places. Lockdown efficacy was assessed by the maximum number of infections recorded during a simulation run. Our analysis shows that lockdown outcomes depend sensitively on the timing of imposition and that it is possible to minimize lockdown durations while limiting case loads to numbers below the hospitalization thresholds. Such timing based effects arise naturally from the non-linear nature of SIR dynamics.

**Notation:** *N* Total Population

*S* Number of susceptible individuals

*I* Number of infected individuals

*R* Number of recovered/removed individuals

*β* Per-individual disease transmission rate

*γ* Recovery rate

*τ* Lockdown start-time

Δ Duration of lockdown

*p* Post-lockdown coefficient

*h* Total number of hospital beds

*ξ* Maximum fraction of infected individuals

*ξ*_0_ Hospitalization threshold

## Introduction

Understanding disease dynamics has assumed vital importance in the midst of the on- going *SARS-CoV2* pandemic. Decision-makers around the world are grappling with the difficult task of weighing the need for lockdown interventions to mitigate disease spread against the negative socioeconomic impact of prolonged disruptions to trade, business and social interactions [Moser and Yared, 2020]. Nowhere else is this issue more critically important than in lower income countries (LICs) and lower middle income countries (LMICs), where policy-makers have to decide between imposing strict lockdowns, which inevitably lead to massive financial difficulties for economically weaker sections of the society[Singh and Neog, 2020], or allow the disease to run unchecked and cause a high number of casualties[Lancet, 2020]. Therefore, modelling strategies that inform decision-making about the optimum timing and duration of lockdown interventions are the need of the hour. However, in order to be effective, models must possess realistic features, which allow for relevant contingencies to be mapped onto model parameters [Diekmann et al., 2000, Singh and Adhikari, 2020, Ray et al., 2020]. One promising approach in that direction is to use Susceptible-Infected-Recovered (SIR) model equations [Hethcote, 2000] in conjunction with empirically obtained age-structured contact matrices to model disease dynamics. Contact matrices may be regarded as the network scaffolding on which SIR dynamics evolve; lockdowns are modelled as scaling operations on contact matrices for home, workplaces, school etc.

Non-linear systems produce differing outputs depending on the timing of application of perturbation[Shulgin et al., 1998, Nie et al., 2012]. For example, neurons in the brain, conceptualized as oscillators that exhibit rhythmic electrical activity, are known to be maximally sensitive to incoming spike currents at specific phases of on-going activity [Canavier, 2006] to generate new action potentials (spike). Similarly, cardiac beats are susceptible to respiratory drive at precise phases of the trajectory [Kralemann et al., 2013]. Researchers use Phase Response Curves(PRCs) to characterize the effects of external perturbations on system dynamics as a function of the phases at which those perturbations are applied [Schultheiss et al., 2011]. We conjecture that such insights can be extended to study the effects of lockdown interventions (perturbations) on the on-going dynamics of disease evolution. Specifically, we hypothesize that the timing of lockdown intervention would considerably impact lockdown outcomes. This approach has the practical benefit of informing public-health planning to best optimize mitigation strategies in order to minimize lockdown durations while avoiding a catastrophic breakdown of healthcare services like the availability of hospital beds for severe cases.

The goal of this article is to identify optimal windows for lockdown interventions subject to a hospitalization threshold (HT) constraint through a numerical analysis of disease dynamics and phase-space analysis. As a first step in that direction, we fit the Covid-19 growth curve in India recorded between 30^*th*^ January, 2020 to 14^*th*^ July, 2020 using a SIR model with relevant model parameters. Next, we use the derived model parameters to explore the parameter space of lockdown timing and duration and produce phase-space trajectories that satisfy relevant criteria.

## Methods

### Model Description

We employ an age-structured SIR model to fit the SARS-Cov2 infection spread on Indian population that can be split into *M* = 16 age-groups ranging from 0-79 years [Desa, 2015]. The contacts among the age groups were classified into four sectors - home (H), school (S), work (W), and other (O) [Singh and Adhikari, 2020]. Dynamics of each age-group is captured by the state variables *S*_*i*_, *I*_*i*_ and *R*_*i*_ which correspond to the number of susceptible, infected and recovered individuals in the *i*^*th*^ age-group (**Figure 1**). The model makes no distinction between recovered or deceased individuals. The dynamics of each age-group is mathematically described by the following ordinary differential equations (ODEs):

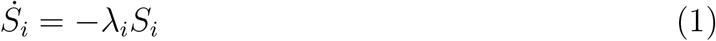

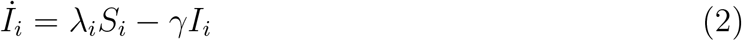

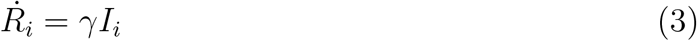

where the effective transmission rate, *λ*_*i*_, is given by :

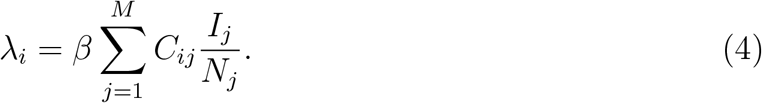

**Figure 1.**
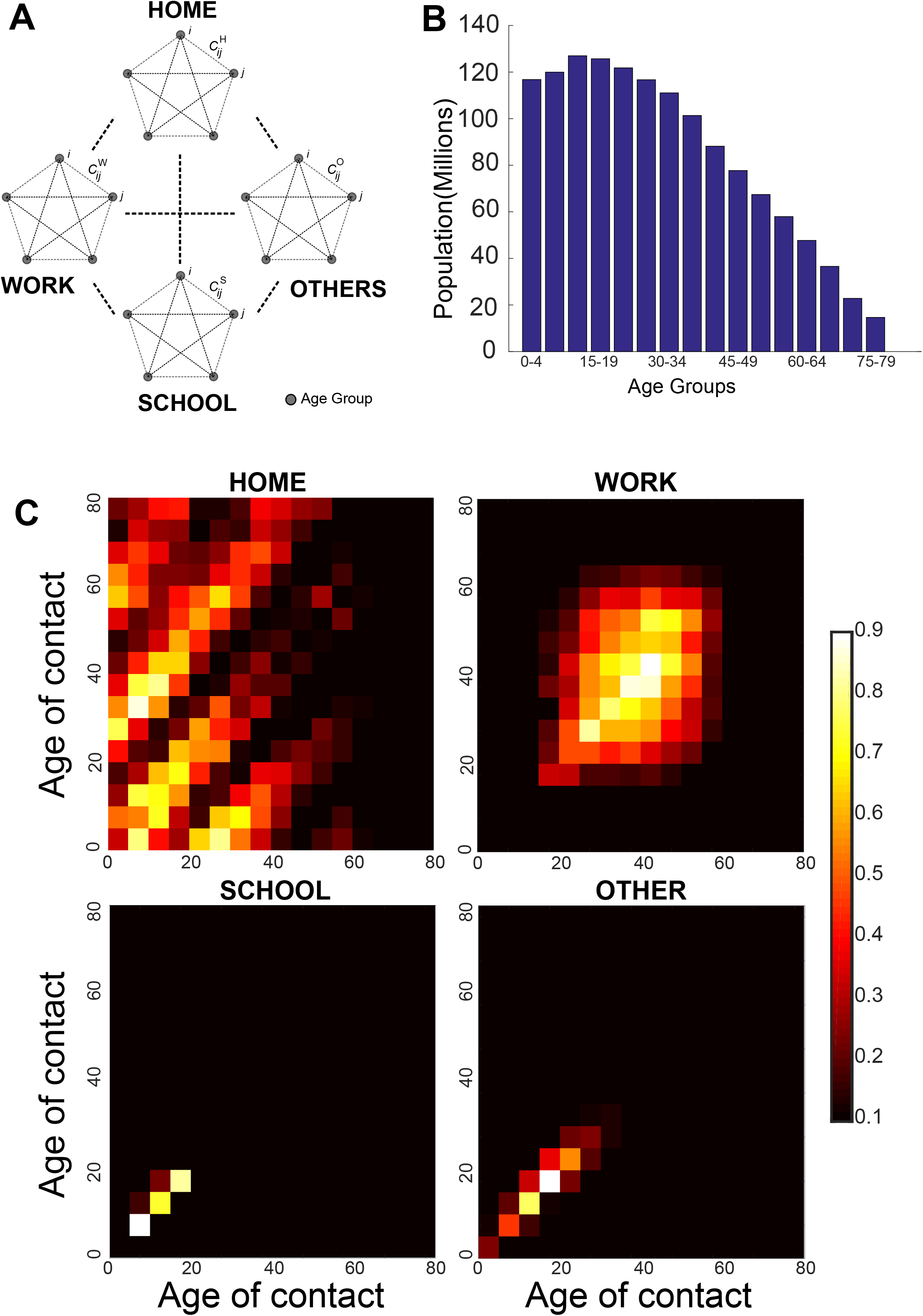
Contact structures and age-demographics of India: **A)** Model schematic, every age group is modelled as a SIR population coupled through contact matrices for home, work, school and other places. **B)** Population split into 16 age groups from 0-79. **C)** contact matrices for Home, work, school and others respectively, plotted as heatmaps. Normalized colorbar indicates magnitude of interaction between age-groups.

*C*_*ij*_ is an element of the contact matrix derived through combination of census surveys and Bayesian imputation [Prem et al., 2017]. *C*_*ij*_ is computed as a linear sum of all contact matrices for *Home, School, work* and *other* places-

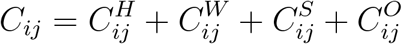

The model ignores birth and death dynamics and therefore,

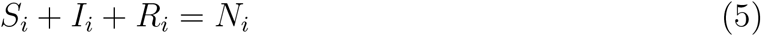

Lockdowns are modelled as interventions that alter the weights of contact matrices for work, school and other places through coefficients *U*_*W*_, *U*_*S*_ and *U*_*O*_ :

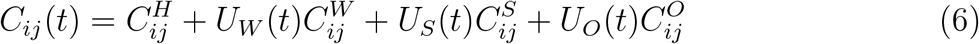

For simplicity we assume *U*_*W*_ (*t*) = *U*_*O*_(*t*) throughout the article. The recovery time is kept at 14 days, throughout (*γ* = 1*/*14). The simulation was initiated with 1 infected individual in the age group 40-44. Euler method was used to integrate the system of 48 ordinary differential equations using an integration time step of 0.1 hr (6 minutes).

To obtain the parameter values to guide further simulations, we fit the simulation against WHO data for the cumulative cases in India till 14th July. The start time for the simulation was kept as 30th January, which also coincided with the start of recorded data. Lockdown imposition was assumed to take place on the 55^*th*^ day from the start of recording. Lockdown was implemented by altering the weightage of contact matrices with time. Prior to lockdown imposition, lockdown parameter coefficients for the *home, work, school*, and *other* connectivity matrix were set to 1. Lockdown was assumed to be lifted on the 115^*th*^ day from the start of the simulation (**Figure 2**). The lockdown state corresponded to reduced coefficients for *work, school* and *other* connectivity matrices(< 1). Post-lockdown coefficients for *work* and *other* connectivity matrices were assumed to be higher than their lockdown values, indicating post-lockdown social distancing measures. The lockdown coefficient for *school* was maintained at 0. The lockdown coefficients and per-individual transmission rate (*β*) were derived from fitting simulation runs to cumulative infection cases recorded till 14^*th*^ July, 2020 [WHO, 2020] (**Figure 2**).

**Figure 2.**
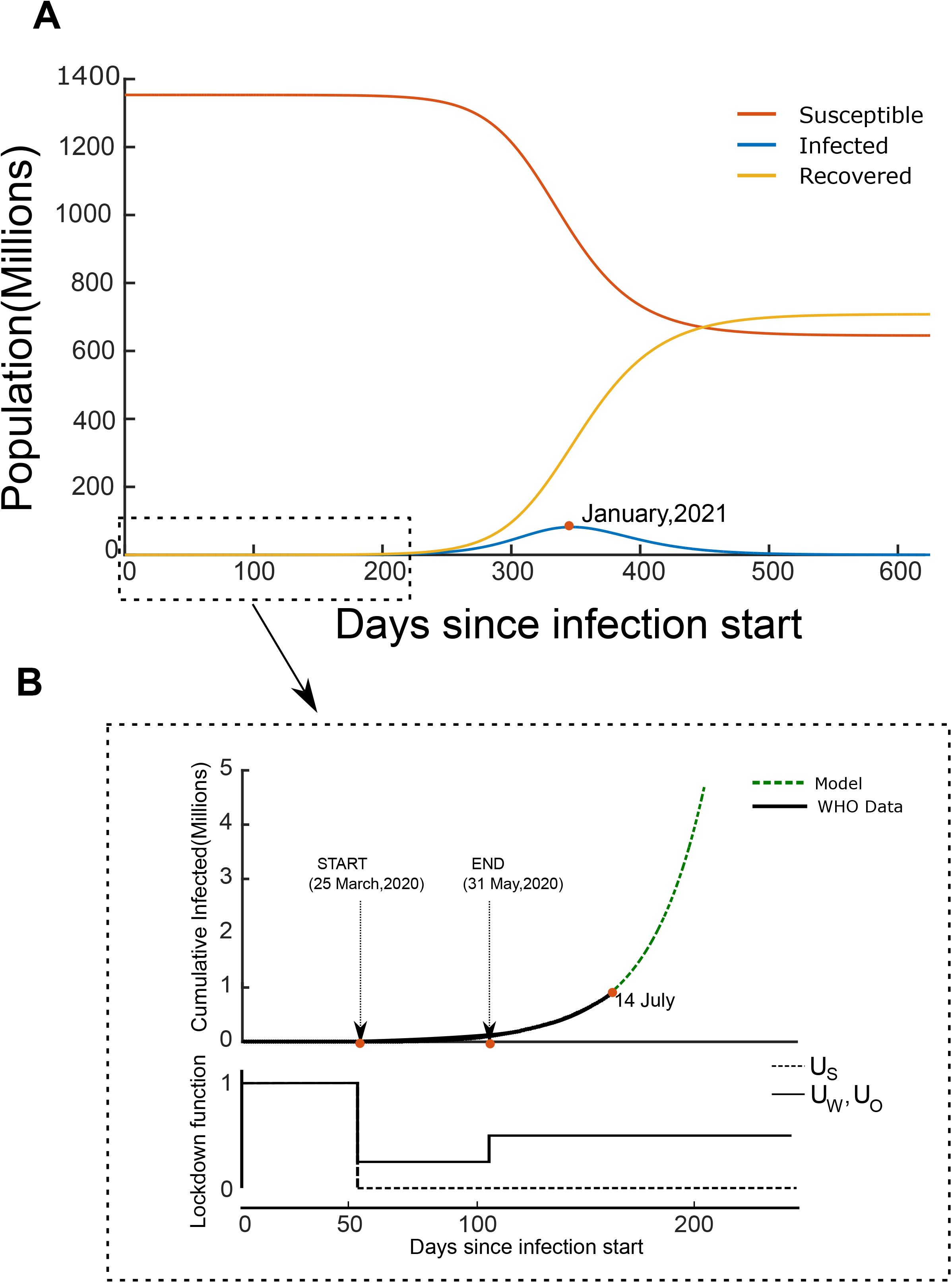
Model fitting from empirical data: **A)** SIR dynamics for model with *β* = 0.013,*U*_*W*_ = *U*_*O*_ = 0.3 during lockdown and *U*_*W*_ = *U*_*O*_ = 0.45 post lockdown. The number of active infected cases peaks on the 346^*th*^ day since the beginning of the lockdown. **B) Inset:**Model fit to cumulative infected cases in India— red indicates WHO data, blue indicates model fit. Lockdown is assumed to have started on 25th March and lifted on 31st May. Model parameters were estimated with data collected till 14th July. **Below)** Model derived lockdown functions for Work, School and Others. Lockdown function for home was considered to be 1 throughout.

A recent report by the Center for Disease Dynamics, Economics and Policy (CDDEP), predicts that at its peak, the infection will affect 100 million people in India out of which 2-4 million (2 − 4%) people will require hospitalization [Klein et al., 2020]. We use this figure of 2 − 4% as an estimate of the maximum hospitalization capacity (*ξ*_0_).

The lockdown start time (*τ*) is varied from 1 to 200 days, and the duration of the lockdown (Δ) is varied from 5 to 200 days. For each simulation, the maximum fraction of infected individuals is computed as:

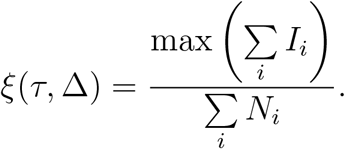

Assuming the hospitalization capacity to be 3% of the infected cases [Klein et al., 2020] and that the number of hospital beds in India (public and private sector combined), *h≈*1.9 million [Kapoor et al., 2020], we determine the hospitalization threshold (HT):

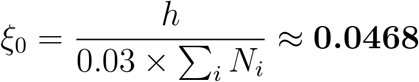

The aim of the study is to identify shortest possible lockdown duration that prevents the number of active cases at any given time from exceeding this value. Formally, this corresponds to the condition:

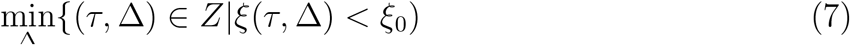

We approach this problem by running the model for ∑_*i*_ *N*_*i*_ = 1353344709 individuals, divided among the 16 age groups for a duration of 25000 hours to ensure that the disease has run its course. The lockdown functions *U*_*W*_ (*t*), *U*_*O*_(*t*), and *U*_*S*_(*t*) are constructed such that its minimum value is 0.3 for *U*_*W*_ and *U*_*O*_, and 0 for *U*_*S*_, consistent with the curve fitting exercise (see Results). The non-zero minimum of the lockdown function captures a *leaky* lockdown scenario, which accounts for sectors where activity cannot be completely arrested, inspite of strict implementation. The lockdown functions are thus defined as:

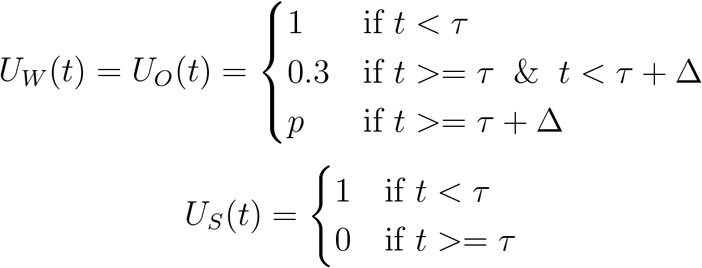

The Δ satisfying (7) can be found by thresholding *ξ* at *ξ*_0_ (**Figure 3**.). In order to illustrate the effect of the post-lockdown coefficients on optimal lockdown strategies, we carry out this exercise for *p* = 0.75 and *p* = 1 (full reopening of the *work* and *other* contact matrix) as well (**Supplementary figures**).

**Figure 3.**
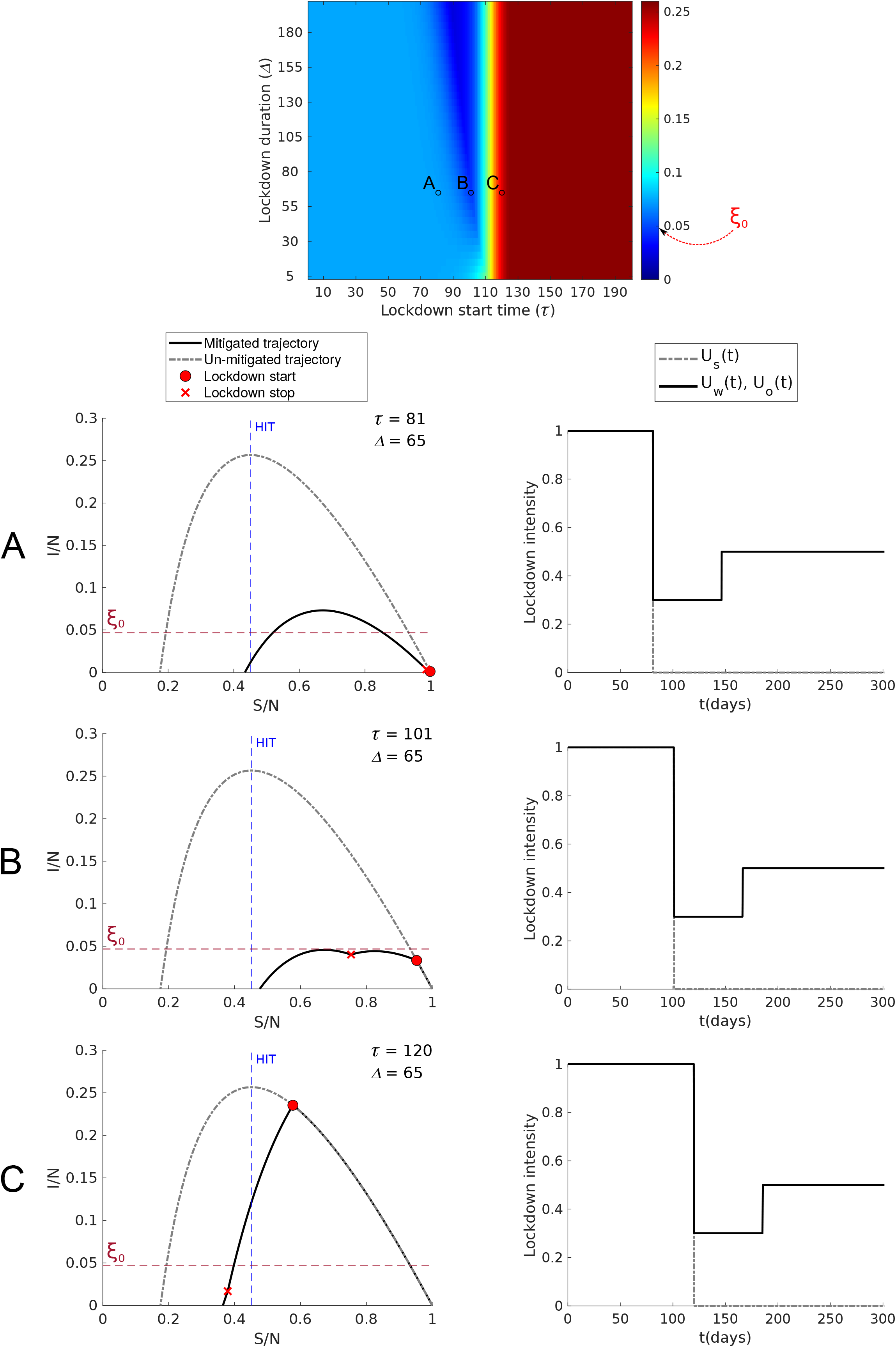
Characterizing lockdowns: Heatmap shows the maximum number of active cases as fraction of total population for model runs with varying lockdown start time(*τ*, days since start of spread) and duration(Δ,in days). **A-C)** indicate the phase trajectories for lockdown strategies marked in the heatmap, along with the corresponding lockdown functions(**right panel**). The red horizontal broken lines(**A-C**) indicates the hospitalization threshold. The blue vertical line marks the Herd-Immunity Threshold (HIT) beyond which the number of active cases are assured to fall. **B)** shows that it is possible to restrict active cases below the hospitalization threshold for the given lockdown parameters. The lockdown function is derived from fitting the model with WHO data.

## Results

### SIR dynamics and lockdown

We observe the characteristic features of the SIR dynamics from our model (**Figure 2**). The number of active cases increase exponentially at first, peak and then fall off. The point at which the unmitigated spread of infection reaches its peak is regarded as the herd-immunity threshold (HIT). The number of recovered individuals increases over time and saturates when the disease spread is terminated.

Lockdown scenarios are modelled by altering lockdown functions *U*_*W*_, *U*_*S*_ and *U*_*O*_ during and after the lockdown. Since schools were shut down during the lockdown and remain shut even 2 months after the formal lifting of nation-wide lockdown, we assume *U*_*S*_ = 0 for all times after lockdown start. We find the best fit to actual data by assuming *β* = 0.013, and *U*_*W*_ = *U*_*O*_ = 0.3 during the lockdown and *U*_*W*_ = *U*_*O*_ = 0.45 after the lockdown (**Figure 2**). According to the model, if the same values of disease infectiousness and *U*_*W*_, *U*_*O*_ and *U*_*S*_ persist as of July 14, the active cases will peak between Dec, 2020 - Jan, 2021, 346 days from the start of the infection.

However, if different phased out lock-downs are performed, the proportion of infected population reaching the hospitalization threshold (HT) can be managed (**Figure 4c,d**). We systematically evaluate the parameter space for optimal lockdowns in the next section.

**Figure 4.**
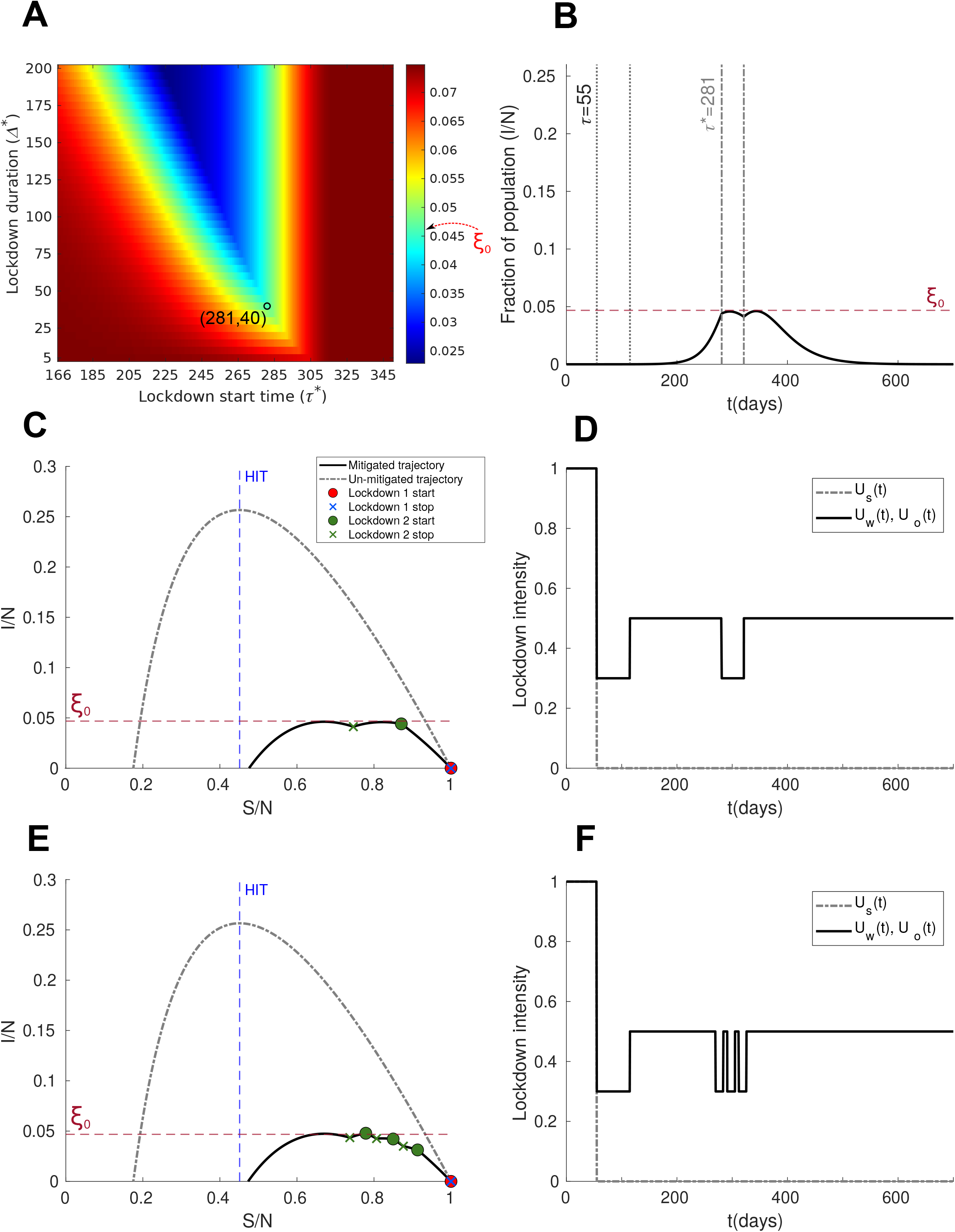
Successive lockdown: A second lockdown around the 281^*st*^ day (*τ*^*\**^) for a duration of 40 days(Δ^*\**^), prevents the number of active cases from breaching the hospitalization threshold. **A)** Heatmap: x-axis indicates lockdown start times from unlock date(166^*th*^ day), y-axis corresponds to lockdown duration. **B)** Trajectory of active cases subject to lockdown function shown in **D). C)** Phase-space evolution of disease spread. **E)-F)** The lockdown is distributed into 3 smaller sub-lockdowns of 14 day each with 7 days between each sub-lockdown.

#### Optimal temporal windows for lockdown

Lockdown parameter space is explored by varying lockdown start time and durations (**Figure 3**). The lockdown functions were assigned values derived from fitting the model against actual data recorded from 30th January to 14th July [WHO, 2020] (**Figure 2**).

The maximum number of active cases computed for each lockdown parameter configuration was compared to numbers that can be maximally catered by the hospital infrastructure and used as a measure of the strain on hospital capacity. As the heatmaps (**Figure 3**) clearly indicate, there exists considerable variability in lockdown outcomes as a function of start times and durations. For example, case A (**Figure 3**) (*τ* = 81, Δ = 65) results in the active infections surpassing the hospitalization threshold (HT). Similarly, case C illustrates that late lockdowns (*τ* = 120, Δ = 65) also result in a violation of the HT. However, same lockdown durations as A & C implemented around the 101^*st*^ day prevent the active cases from breaching the HT. This phenomenon, which results from the non-linearity inherent in SIR dynamics, is easily visualized by plotting disease trajectory in the phase space(S vs I) (**Figure 3**).

The optimal lockdown window that prevents a breach of the HT can also be found in the cases where post-lockdown values of *U*_*W*_ and *U*_*O*_ are set to 0.75 or 1 (**Supplementary figures**). However, in these cases, the minimum duration of a favourable lockdown strategy is considerably greater (100 days and 135 days for *U*_*W*_ = *U*_*O*_ = 0.75 and 1 respectively).

We also evaluate the outcomes of a possible successive lockdown to restrict *ξ* below *ξ*_0_ (**Figure 4**). Our results show that a second, 40 day nation-wide lockdown can potentially contain active cases below the HT, if executed around the 281^*st*^ day (October, 2020) (**Figure 4**). **Figure 4e**,**f** also show that it may be possible to distribute the *∼*40 day lockdown into 3 smaller sub-lockdowns, each 14 day long with 7 days relaxation between each. **Figure 4e** shows that splitting the lockdown leads to a breach of HT, while permitting week-long relaxation windows between the sub-lockdowns. We have explored further by varying the inter-lockdown periods from 7 days to 35 days, all of which led to crossing the HT by a considerable amount.

## Discussion

In response to the global SARS-CoV2 pandemic, the Government of India imposed a 4 phase nation-wide lockdown that lasted from 25^*th*^ March, 2020 to 31^*st*^ May, 2020. This was followed by 2 phases of *unlocks*, with the second unlock phase still underway at the time of writing. While being a necessary step to ensure disease mitigation, the lockdown brought tremendous hardship to the economy, causing disruption in livelihoods, supply-chains and stalling growth [Dev et al., 2020, Buheji et al., 2020]. These factors ultimately led to calls for lifting of the lockdown.

The main focus of the article was to estimate optimal lockdown windows such that, (*i*) the maximum number of active cases in the resulting dynamics remain lower than the total hospitalization capacity, and (*ii*) condition (*i*) is satisfied for the smallest possible lockdown duration. As argued in the introduction, such a solution ensures medical assistance to the most severe cases, while keeping the economic fallout of a protracted lockdown to a minimum. Also it should be emphasized in this context that even though we have used empirical data to tune our model parameters, our model by no means aim to predict the exact unfolding of events in future. Rather, our study demonstrates that the concept of optimal lockdowns exist because of the complex dynamical behavior of the SARS-CoV2 infection spread. Subsequent studies are necessary to predict the nature of lockdown functions that will work in a specific population, that will obviously be dependent on the contact matrices. One clear departure from the Singh and Adhikari [Singh and Adhikari, 2020] study on the Indian population is a clear demonstration that an early lockdown doesn’t seem to have a very high success probability because the social costs are too high, and premature withdrawal will lead to increase of infection again. India is credited with imposing one of the most stringent lockdowns as measured by the lockdown stringency index devised by the Oxford University [Hale et al., 2020]. Inspite of this, the lockdown period coincided with an increase in overall case loads [Pulla, 2020] (**Figure 2**). Therefore, we regarded lockdown intensity as an important criteria that demands explicit factoring into any realistic model to assess lockdown scenarios.

In order to keep the model as simple as possible, we restricted ourselves to one-shot, square-wave like lockdown functions, such that the only relevant lockdown parameters were the start-time, duration and intensity of the lockdown. We use an age-structured SIR model to fit actual data (cases and lockdown times) in order to estimate relevant parameters— per-individual transmission rate(*β*) and lockdown intensity during and after the lockdown implementation (**Figure 2**). Even a simple SIR model with realistic age-based compartmentalization gives rise to rich dynamics due to the non-linearity inherent in equations (1) and (2). This richness is manifested in the sensitive dependence of outcomes of mitigation strategies on the timing and duration of the lockdown. This effect can be intuitively understood as arising from the interaction of lockdown measures and herd-immunity thresholds (HITs) (**Figure 3**). Early lockdowns doubtless lead to immediate cessation of disease spread. However, early lockdowns make a second wave of infection inevitable because the number of susceptible individuals in the population after the lockdown remains comparable to the number of susceptible individuals at the start of the pandemic, far from the HIT of the system. In this scenario, infections climb again once the lockdown is lifted [Rachel, 2020, Malani et al., 2020]. Therefore, such lockdowns do not lead to good long-term outcomes as the second wave of the infection easily takes the number of active cases past the hospitalization threshold (HT).

On the other extreme, late lockdowns do not help either as the worst phase of the epidemic is already over at this point and the HTs have already been breached. Therefore, it is evident that the most suitable solutions that prevent medical infrastructure overload, while reducing the lockdown duration, lie somewhere in the middle. Successful lockdown strategies leverage the mitigatory effects of lockdowns along with inherent HITs (beyond which the number of infected individuals strictly decrease). As argued in the introduction, such effects can be likened to phase resetting curves in neuroscience, where this concept is used to estimate the probability of neural spiking as a function of the timing of an external perturbation [Canavier, 2006, Schultheiss et al., 2011].

One strategy being increasingly administered by several local authorities is a short lock-down, instead of a long one to avoid the economic costs. Our analysis based on the SIR model reveals that such a step may require at least a minimum of 40 days of lockdown (**Figure 4**). This estimate is undoubtedly achieved, taking into account the country-wide contact matrices of the Indian population in picture. Nonetheless, a minimum lockdown requirement can be worked out following our approach, if contact matrices are available for any arbitrary population size for which lockdown is being implemented.

### Assumptions and Limitations

We conclude by laying out the inherent assumptions and possible limitations of the model presented here. Most importantly, our model falls in the class of conceptual models to understand which broad lockdown strategy is effective, not necessarily a detailed plan for implementing the lockdown across diverse socio-culturally bound populations. For example, our model ignores temporal delays that are likely to exist between lockdown announcement and effective implementation. We think that such delays would not amount to substantial differences in disease dynamics as it is possible that the effects of the delay are offset by a gradual reduction in social mobility preceding the actual lockdown announcement. Additionally, this assumption simplifies the model considerably. Certainly country specific delays would change the predictions from the model.

Second, by assuming a single value for *ξ*_0_, the model implicitly assumes spatial homogeneity in the distribution of medical infrastructure across the country. However, both disease burden and medical infrastructure is known to be heterogeneously distributed across the country [Klein et al., 2020]. Detailed modelling at the level of individual states and districts could potentially solve the problem but at considerable computational costs. We decided to favour model simplicity since our focus was on assessing optimal lockdown scenarios.

Third, it is well-known that the number of reported cases is a function of the number of tests conducted. Our model does not account for fluctuations in the rate of testing that may influence the total number of infected individuals at any given time. Further we don’t account for the inherent birth and death rates of the population during the course of the infection.

Existence of optimal lockdown windows depends upon the choice of the quantity that we are aiming to optimize. The current analysis places a premium on the maximum fraction of infected individuals (*ξ*) which serves as a measure of strain on the medical infrastructure at any given point of time. Indeed, recent events have demonstrated the need for *curve flattening* in order to de-congest hospitals and reduce the burden on health-care professionals. However, it may be useful to frame the problem within a formal optimization framework to optimize quantities derived from other economic considerations [Rachel, 2020].

## Data Availability

All data used here is freely available-
WHO coronavirus dashboard
Population pyramid

https://covid19.who.int/?gclid=EAIaIQobChMIjNeQ4_X26gIVTg4rCh2yMQQAEAAYASAAEgJqDvD_BwE

https://www.populationpyramid.net/world/2019/

## Acknowledgements

This work was supported by NBRC core funds.

## Abbreviations

SIR: Susceptible, Infected, Recovered/Removed
HIT: Herd Immunity Threshold
HT: Hospitalization Threshold

---

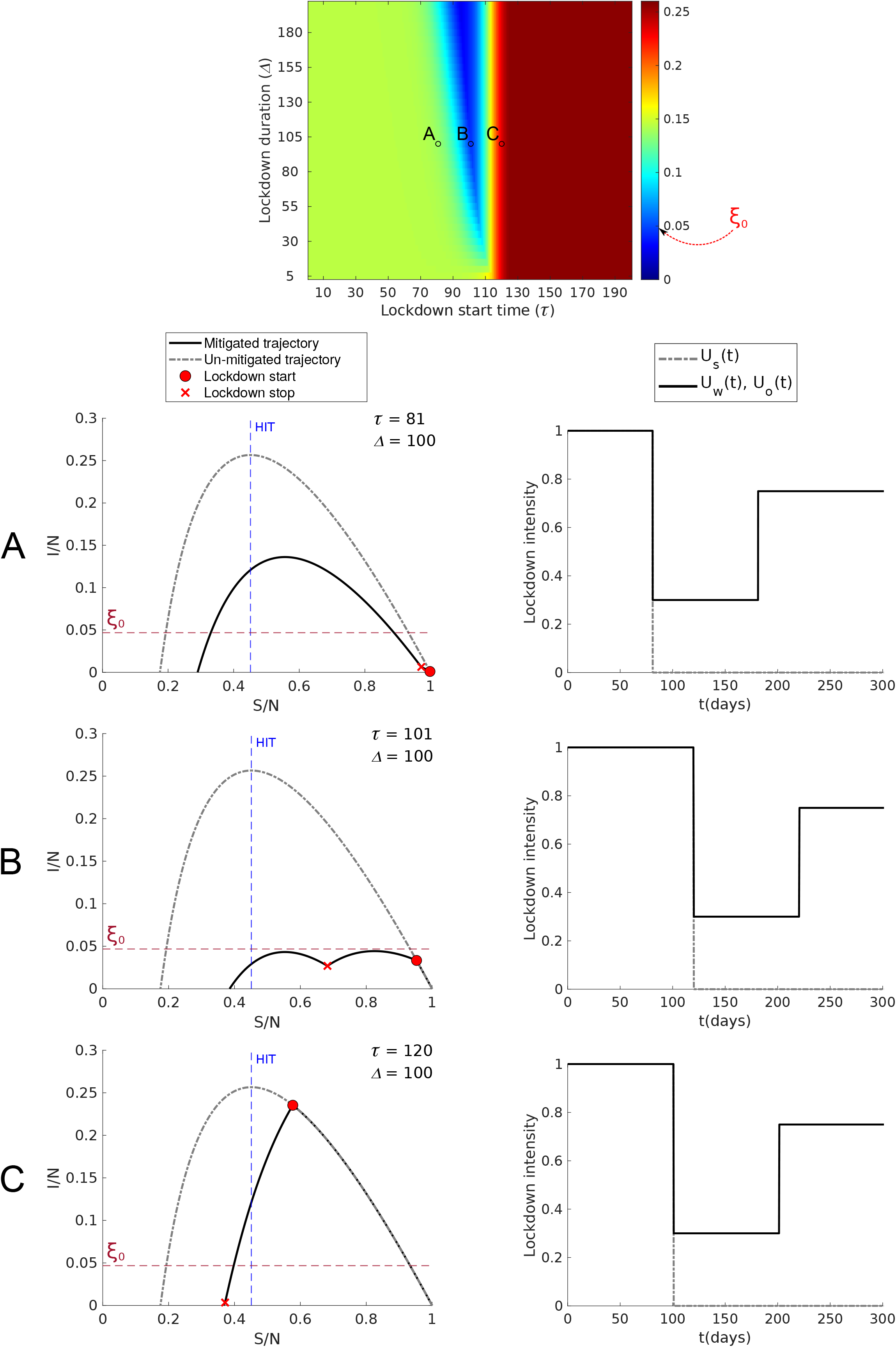

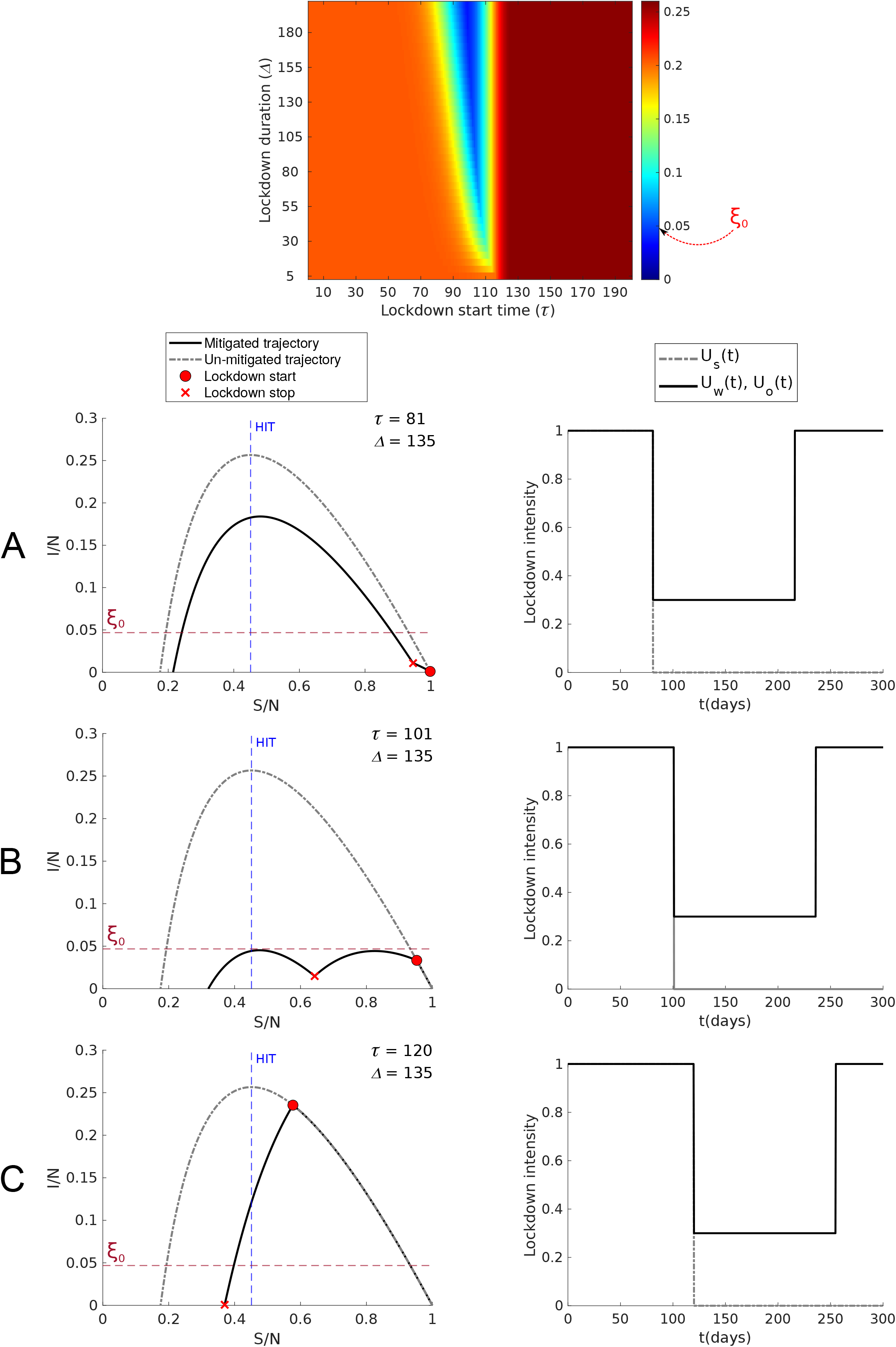

